# Gut microbiome composition is predictive of incident type 2 diabetes

**DOI:** 10.1101/2021.11.10.21266163

**Authors:** Matti O. Ruuskanen, Pande P. Erawijantari, Aki S. Havulinna, Yang Liu, Guillaume Méric, Michael Inouye, Pekka Jousilahti, Veikko Salomaa, Mohit Jain, Rob Knight, Leo Lahti, Teemu J. Niiranen

**Affiliations:** Department of Computing, University of Turku, Turku, Finland; Department of Public Health and Welfare, Finnish Institute for Health and Welfare, Helsinki, Finland; Institute for Molecular Medicine Finland, FIMM - HiLIFE, Helsinki, Finland; Cambridge Baker Systems Genomics Initiative, Baker Heart and Diabetes Institute, Melbourne, Victoria, Australia; Department of Clinical Pathology, Melbourne Medical School, The University of Melbourne, Melbourne, Victoria, Australia; Department of Infectious Diseases, Central Clinical School, Monash University, Melbourne, Victoria, Australia; Department of Public Health and Primary Care, Cambridge University, Cambridge, United Kingdom; Department of Medicine, University of California, San Diego, La Jolla, California, USA; Department of Pharmacology, University of California, San Diego, La Jolla, California, USA; Jacobs School of Engineering, University of California, San Diego, La Jolla, California, USA; Center for Microbiome Innovation, University of California San Diego, La Jolla, California, USA; Department of Pediatrics, School of Medicine, University of California San Diego, La Jolla, California, USA; Department of Computer Science & Engineering, University of California San Diego, La Jolla, California, USA; Division of Medicine, Turku University Hospital, Turku, Finland

**Keywords:** Metagenomics, human gut, type 2 diabetes, survival analysis, population sample

## Abstract

**OBJECTIVE:** To examine the previously unknown long-term association between gut microbiome composition and incident type 2 diabetes in a representative population cohort.

**RESEARCH DESIGN AND METHODS:** We collected fecal samples of 5 572 Finns (mean age 48.7 years, 54.1% women) in 2002 who were followed up for incident type 2 diabetes until Dec 31^st^, 2017. The samples were sequenced using shotgun metagenomics. We examined associations between gut microbiome compositions and incident diabetes using multivariable-adjusted Cox regression models. We first used the Eastern Finland sub-population to obtain initial findings and validated these in the Western Finland sub-population.

**RESULTS:** Altogether 432 cases of incident diabetes occurred over the median follow-up of 15.8 years. We detected 4 species and 2 clusters consistently associated with incident diabetes in the validation models. These 4 species were *Clostridium citroniae* (HR, 1.21; 95% CI, 1.04-1.42), *C. bolteae* (HR, 1.20; 95% CI, 1.04-1.39), *Tyzzerella nexilis* (HR, 1.17; 95% CI, 1.01-1.36), and *Ruminococcus gnavus* (HR = 1.17; 95% CI, 1.01-1.36). The positively associated clusters, cluster 1 (HR, 1.18; 95% CI, 1.02-1.38) and cluster 5 (HR, 1.18; 95% CI, 1.02-1.36), mostly consisted of these same species.

**CONCLUSIONS:** We observed robust species-level taxonomic features predictive of incident type 2 diabetes over a long-term follow-up. These findings build on and extend previous mainly cross-sectional evidence and further support links between dietary habits, metabolic diseases, and type 2 diabetes that are modulated by the gut microbiome. The gut microbiome could potentially be used to improve risk prediction and to uncover novel therapeutic targets for diabetes.

---

The roles of host genetics and environmental factors in the pathogenesis of type 2 diabetes have been widely studied (1,2). Recently, several studies have reported a link between gut microbiome composition and type 2 diabetes (3–5). These associations could involve several mechanisms, such as modulation of inflammation, increased gut permeability, interactions with dietary constituents, glucose and lipid metabolism, insulin sensitivity, and effects on overall energy homeostasis of the host (5). Specifically, type 2 diabetes has been reported to be associated with lower relative abundances of butyrate-producing microbes and increases in various opportunistic pathogens (4,6).

Most prior studies on the association between gut microbiome and type 2 diabetes have been limited by their cross-sectional designs (3,5). While these studies have begun to elucidate the role of the gut microbiome in type 2 diabetes pathogenesis, they are subject to selection bias and have not included prospective data on incident diabetes. As a result, such analyses provide limited information on how the gut microbiome could be used in diabetes risk prediction. Prospective studies have thus far been conducted rarely, with short-term follow-ups (7), or only in the context of diurnal oscillation of gut bacteria (8). In addition, growing evidence indicates that some previous results from cross-sectional studies might be confounded by the use of anti-diabetic drugs that influence gut microbiome composition, such as metformin (9,10).

Here, we analyzed the long-term association between gut microbiome composition and incident type 2 diabetes in a well-phenotyped and representative Finnish population sample (n = 5 572). The follow-up spanned 16 years after sampling (11). Notably, participants with prevalent diabetes at baseline, including those taking anti-diabetic drugs such as metformin, were excluded from our study. The FINRISK 2002 cohort features participants both from Eastern and Western Finland with differences in genetics, lifestyles, morbidity, and mortality rates (12). To improve the robustness of our results, we performed feature selection separately in data from Eastern Finland and evaluated the findings in participants from Western Finland to establish robust microbial signals, predictive of incident type 2 diabetes.

## Research Design and Methods

The FINRISK study has been conducted in Finland to investigate risk factors for cardiovascular disease every 5 years since 1972 (11). In 2002, the study included participants from six areas: North Karelia; Northern Savo; Oulu; Lapland; Turku and Loimaa; and Helsinki and Vantaa.

These areas can be geographically divided roughly to Western Finland (Turku and Loimaa; Helsinki and Vantaa), and Eastern Finland (North Karelia; Northern Savo; Oulu; Lapland). A random sample stratified (by sex and 10-year age groups) among the populations aged 24 - 74 years was taken in each study area. Out of the 13 498 invitees, 8 783 participated in the study. Out of these participants, 7 231 donated fecal samples. In the current study, we excluded individuals with ≥ 1 exclusion factors: prevalent diabetes (n = 698), pregnancy (n = 40), < 50 000 mapped reads (n = 20) or antibiotic use in the past 6 months (n = 907). After these exclusions, samples from 5 572 participants were eligible for this study.

The health status of the participants was assessed at baseline in 2002 (11). The physical examination and blood sampling were performed at local health centres or other survey sites by trained nurses. Data were collected for physiological measures, biomarkers, and dietary, demographic, and lifestyle factors (11). Willing participants were given a stool sampling kit with detailed instructions. Samples were mailed overnight under Finnish winter conditions to the laboratory, where they were immediately stored at −20°C. The samples were stored unthawed until 2017, when they were shipped to the University of California San Diego for sequencing. The Coordinating Ethics Committee of the Helsinki University Hospital District approved the study protocol for FINRISK 2002 (Ref. 558/E3/2001), and all participants have given their written informed consent.

National healthcare registers in Finland enable combining of the data in FINRISK with subsequent in- and outpatient disease diagnoses and drug prescriptions based on individual personal identity codes. Prevalent and incident diabetes (at baseline) were defined based on ICD10 codes E10-E14, ICD9 code 250, or ICD8 code 250 in the nationwide Care Register for Health Care (HILMO). In addition, prevalent diabetes was based on ≥ 3 drug purchases with ATC drug code A10 in the nationwide Drug Reimbursement Register prior to baseline. This drug code (and thus, the exclusion) includes metformin which is widely reported to alter the gut microbiota (10). The record data was amended with the patient’s self-report, a measured fasting plasma glucose ≥ 7.0 mmol/L, a 2h oral glucose tolerance test plasma glucose ≥ 11.1 mmol/L, or HbA1c ≥ 48 mmol/mol at baseline examination. Glucose tolerance test was only available for 3 378 participants and HbA1c for 4 096 participants (out of 5 572). The participants were followed through December 31^st^, 2017.

Earth Microbiome Project protocols were utilized for DNA extraction using the MagAttract PowerSoil DNA kit (Qiagen), as described previously (13). Library generation was performed with a miniaturized version of the Kapa HyperPlus Illumina-compatible library prep kit (Kapa Biosystems) (14). Echo 550 acoustic liquid handling robot (Labcyte Inc.) was used to normalize DNA extracts to 5 ng total input per sample. 1/10 scale enzymatic fragmentation, end-repair, and adapter-ligation reactions were performed with a Mosquito HV liquid-handling robot (TTP Labtech Inc.). Sequencing adapters were based on the iTru protocol (15), where ligation of short universal adapter stubs is followed by addition of sample-specific barcoded sequences in a subsequent PCR step. PicoGreen assay was used to quantify amplified and barcoded libraries, which were pooled in approximately equimolar ratios before being sequenced on an Illumina HiSeq 4000 instrument. An average read count of 900 000 reads per sample was achieved with this protocol. Atropos was used for quality trimming of the sequences and removal of sequencing adapters (16). Bowtie2 (17) was used to remove host reads by mapping them against the human genome assembly GRCh38. SHOGUN v1.0.5 (18) was utilized to assign taxonomy to the reads using NCBI RefSeq version 82 (May 8, 2017), which contains complete bacterial, archaeal, and viral genomes, together with plasmid sequences.

All statistical analyses were performed with R 3.6.1 (19). The data was first divided into participants from Eastern Finland (3 871 participants) and Western Finland (1 701 participants). These subpopulations were selected because of their well-known differences in genetic backgrounds, lifestyles and mortality rates (12). Because of the larger number of participants in Eastern Finland, we used this data set to discover associations, followed by validation of the findings with Western Finland data. Alpha diversity of the microbiomes was assessed with raw counts per taxa and Shannon diversity. Beta diversity was calculated separately in the data from Eastern and Western Finland sub-populations by applying a centered log-ratio (CLR) transformation on the taxa counts followed by principal component analysis. Rare taxa were filtered out in Eastern Finland data with the cutoffs set at detection > 0.01% and prevalence > 1% of raw (untransformed) mapped reads. Taxa were then subset to this filtered set (119 taxa) and CLR-transformed in all of the data.

Cox proportional hazards regression models for survival time were first constructed solely in data from Eastern Finland with the R package ‘survival’ 3.2.11 (20). Models were constructed for 1) observed counts (total number of raw taxa matches), 2) Shannon diversity 3) the first 10 PC axes (10 separate models), and 4) relative taxa abundances (119 separate models). Each model was adjusted for baseline age, body mass index, sex, systolic blood pressure, non-high-density lipoprotein (non-HDL) cholesterol, triglycerides, and current smoking of the participants. Features significantly associated with incident type 2 diabetes were filtered at alpha level p < 0.05 after applying a Benjamini-Hochberg correction. Instead of correlation, we analyzed the compositionally valid measure, proportionality (ρ), between the significantly associated taxa using the R package ‘propr’ 4.2.6 (21). The taxa were then clustered based on proportionality with Ward’s minimum variance method and the optimal number of clusters was defined with Kelley-Gardner-Sutcliffe penalty function in the R package ‘maptree’ 1.4.7 (22). Heatmaps of the proportionality between taxa, and associated clusters and hazard ratios were visualized with the R package ‘ComplexHeatmap’ 2.7.11 (23). Relative abundances of the clusters were calculated by combining and CLR-transforming the raw counts of the taxa within the data from Eastern Finland.

Following the screening and selection of significant features in the data from Eastern Finland, models were constructed identically and separately in Western Finland. A feature selected in data from Eastern Finland was considered to be robustly predictive of incident type 2 diabetes in the Western sub-population if the 95% confidence interval (CI) of its HR did not overlap 1.0 (unadjusted p < 0.05). Finally, Kaplan-Meier curves were constructed for relative abundance quantiles of these robustly predictive features in data from Western Finland with the R package ‘rms’ 6.2.0 (24).

The source code used to analyze the data and produce our results is included in the Supplementary Information. Due to sensitive health information of individuals, the datasets analyzed during the current study are not public, but are available based on a written application to the THL Biobank as instructed in: https://thl.fi/en/web/thl-biobank/for-researchers

## Results

The characteristics of the study participants are reported in **Table 1**. A total of 432 (7.8%) participants were diagnosed with type 2 diabetes over a median follow-up of 15.8 years.

**Table 1.** Baseline statistics of the participants in FINRISK 2002 after exclusions.

| Variable | Incident type 2 diabetes |  |  |  | Geographical area |  |  |
| --- | --- | --- | --- | --- | --- | --- | --- |
|  | Total | Yes | No | P-value | Eastern Finland | Western Finland | P-value |
| **Number of participants (%)** | 5 572 | 432 (7.8) | 5 140 (92.3) | - | 3 871 (69.5) | 1 701 (30.5) | - |
| **Women (%)** | 3 013 (54.1) | 218 (50.5) | 2 795 (54.4) | 0.12 | 2 074 (53.6) | 939 (55.2) | 0.72 |
| **From Eastern Finland (%)** | 3 871 (69.5) | 293 (67.8) | 3 578 (69.6) | 0.45 | - | - | - |
| **With Incident type 2 diabetes (%)** | 432 (7.8) | - | - | - | 293 (7.6) | 139 (8.2) | 0.45 |
| **Baseline age (years)** | 48.7±12.8 | 52.8±10.6 | 48.4±13.0 | $1.0 \times 10^{-11}$ | 48.7±12.9 | 48.7±12.8 | 0.99 |
| **BMI (kg/m<sup>2</sup>)** | 26.6±4.4 | 30.7±5.2 | 26.3±4.2 | $1.2 \times 10^{-71}$ | 26.8±4.4 | 26.2±4.4 | $3.4 \times 10^{-7}$ |
| **Systolic blood pressure (mm Hg)** | 135.2±19.8 | 141.7±20.4 | 134.6±19.6 | $6.4 \times 10^{-13}$ | 135.9±20 | 133.5±19.2 | $2.8 \times 10^{-5}$ |
| **non-HDL cholesterol (mmol/L)** | 4.1±1.1 | 4.5±1.3 | 4.0±1.1 | $1.1 \times 10^{-14}$ | 4.1±1.1 | 4.0±1.1 | $1.2 \times 10^{-7}$ |
| **Glucose tolerance test, 0 hr (mmol/L)** | 5.7±0.5 | 6.1±0.5 | 5.7±0.5 | $9.2 \times 10^{-34}$ | 5.7±0.5 | 5.7±0.5 | $1.1 \times 10^{-3}$ |
| **Glucose tolerance test, 2 hrs (mmol/L)** | 6.3±1.7 | 7.5±1.9 | 6.2±1.6 | $8.3 \times 10^{-24}$ | 6.3±1.7 | 6.4±1.7 | 0.03 |
| **Hemoglobin A1C (mmol/mol)** | 35.8±3.6 | 38.4±3.8 | 35.5±3.5 | $4.9 \times 10^{-29}$ | 35.6±3.8 | 36.2±3.1 | $7.9 \times 10^{-5}$ |
| **Triglycerides (mmol/L)** | 1.4±0.9 | 1.9±1.3 | 1.3±0.8 | $3.4 \times 10^{-38}$ | 1.4±0.9 | 1.4±0.9 | 0.11 |
| **Current smoking (%)** | 1327 (23.8) | 111 (25.7) | 1216 (23.7) | 0.38 | 914 (23.6) | 413 (24.3) | 0.63 |

- Mann-Whitney U test was used for numerical data and Fisher's exact test for categorical data
- Numerical data are shown as mean ± standard deviation
- Categorical data are shown as number of participants in the indicated category and percentage of the total

In the data from Eastern Finland, out of the 119 taxa remaining after filtering, the relative abundances of 18 were significantly associated with incident type 2 diabetes (adjusted p < 0.05; **Figures 1, 2, Table S1**). 15 taxa had positive associations with incident type 2 diabetes and 3 taxa were negatively associated. Most of the positively associated taxa were from the family *Lachnospiraceae*, with several representatives of genus *Clostridium*. Two of the three negatively associated taxa were from genus *Alistipes*. Alpha diversity was not significantly associated with incident type 2 diabetes (adjusted p > 0.05). In the beta diversity analysis, the first PC axis had a significant association (HR, 0.82; 95% CI 0.69-0.88; adjusted p = 0.01). Significantly associated taxa could be grouped by proportional abundance into 5 clusters (**Figure 1**). 4 taxa and 2 clusters were positively associated with incident type 2 diabetes in the Western Finland sub-population (**Figure 2, Table S1**). These taxa were *Clostridium citroniae* (Eastern Finland HR, 1.21; 95% CI, 1.09-1.35; Western Finland HR, 1.21; 95% CI, 1.04-1.42; unadjusted p = 0.02), *C. bolteae* (Eastern Finland HR, 1.18; 95% CI, 1.07-1.30; Western Finland HR, 1.20; 95% CI, 1.04-1.39; unadjusted p = 0.01), *Tyzzerella nexilis* (Eastern Finland HR, 1.16; 95% CI, 1.05-1.29; Western Finland HR, 1.17; 95% CI 1.01-1.36; unadjusted p = 0.03), and *Ruminococcus gnavus* (Eastern Finland HR, 1.18; 95% CI, 1.06-1.30; Western Finland HR, 1.17; 95% CI, 1.01-1.36; unadjusted p = 0.04). The directions of these associations were the same as those in Eastern Finland.

**Figure 1.**
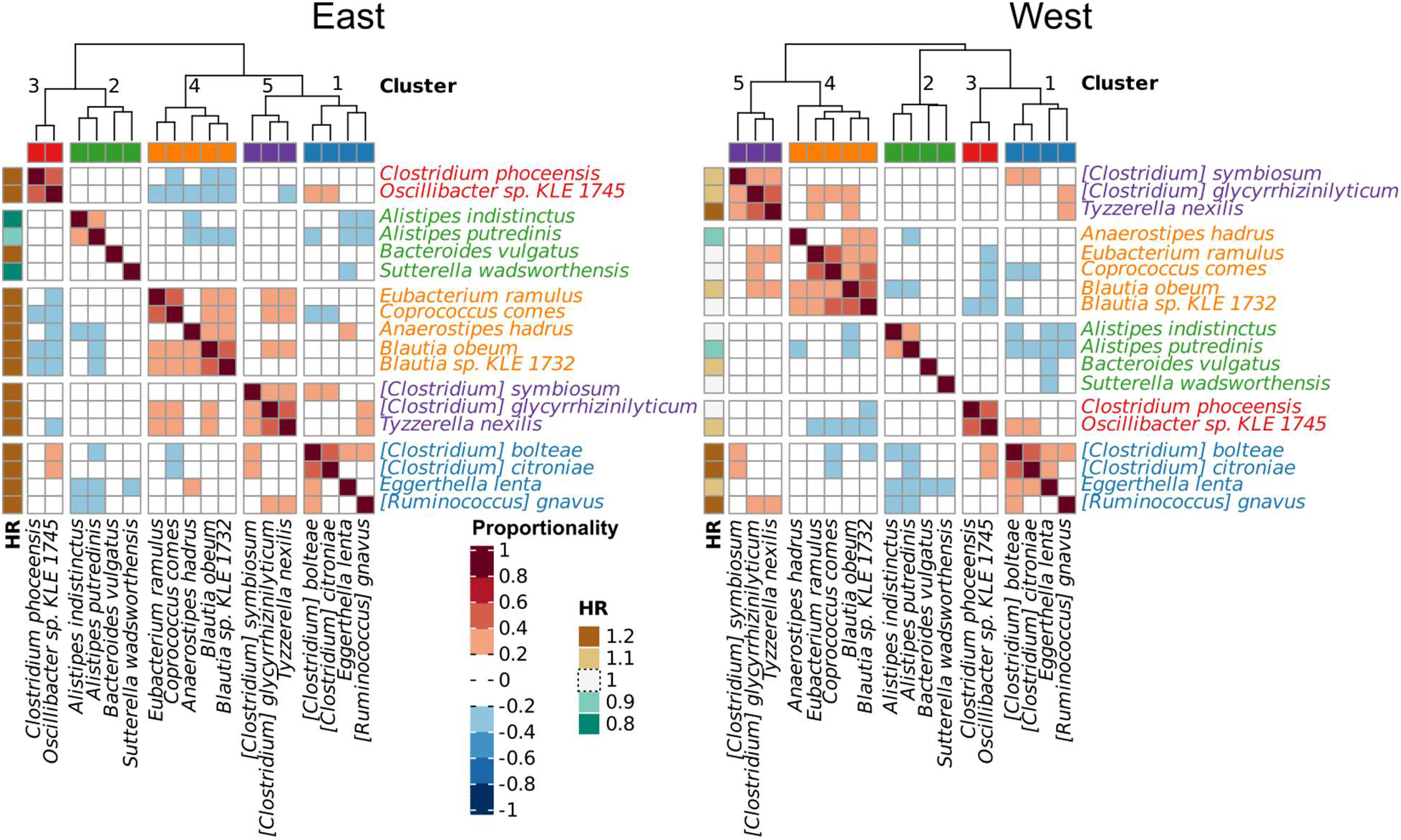
Proportionality between bacterial taxa significantly associated with incident type 2 diabetes in Eastern Finland and Western Finland. Annotated hazard ratios and clustering of the taxa have been calculated separately in both data. Because of identical cluster membership of the taxa, the cluster numbers and their annotation have been harmonized.

**Figure 2.**
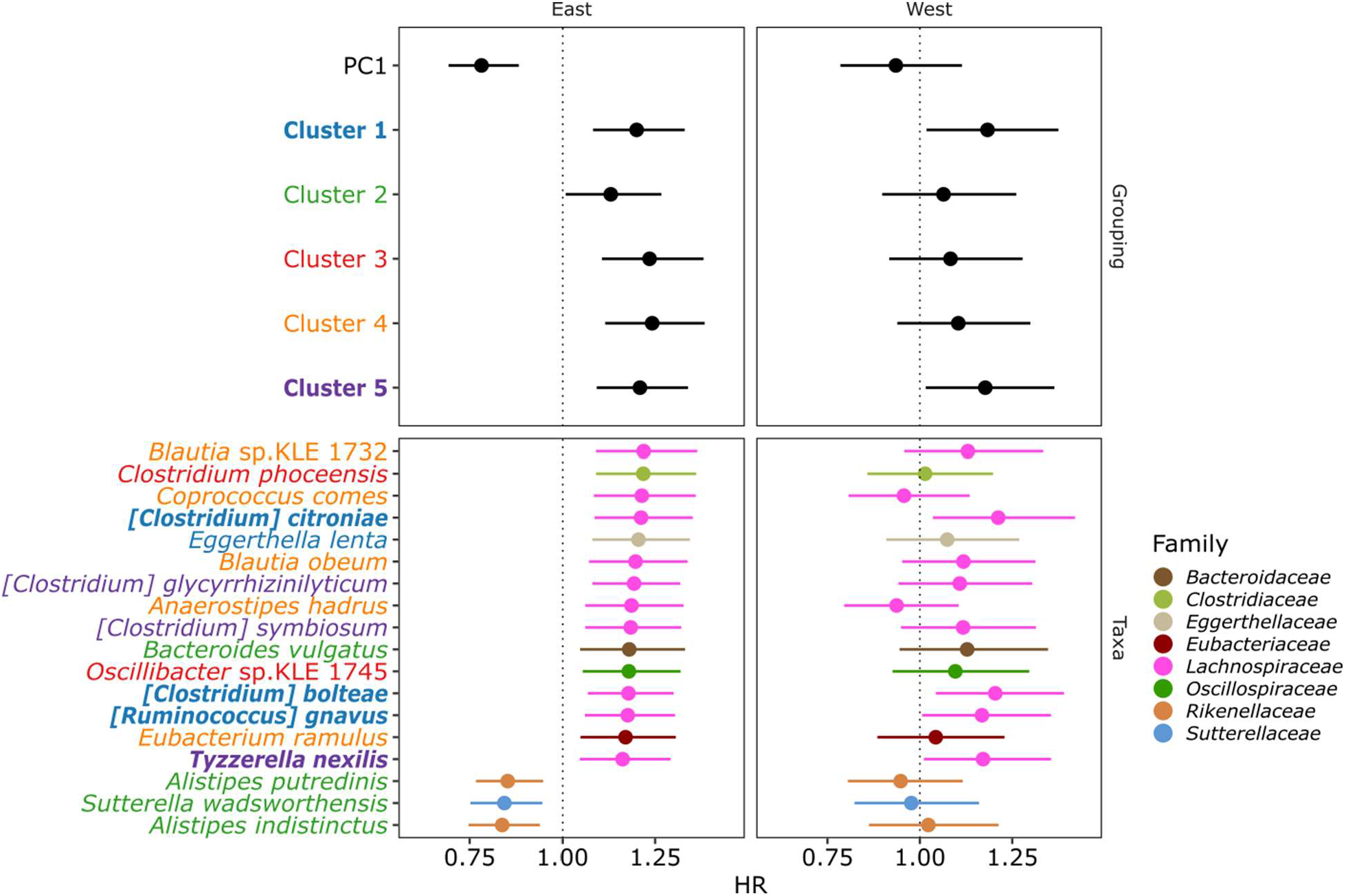
Comparison of hazard ratios between models for the selected features in Eastern and Western Finland data. Features with significant associations in the validation (Western Finland) data are indicated in bold and the taxa colors show their membership in a cluster. The information in this figure can be found in numeric format in **Table S1**.

Clustering the 18 selected taxa by proportional abundance separately in each sub-population data also produced clusters with identical taxa membership (**Figure 1**). Three of the associated taxa, *C. citroniae, C. boltae* and *R. gnavus*, were grouped in cluster 1 (Western Finland HR, 1.18; 95% CI 1.02-1.38; unadjusted p = 0.03) with one additional taxon in the cluster, *E. lenta*, which was not associated with type 2 diabetes in Western Finland data as an individual predictor. *T. nexilis* was grouped in cluster 5 (Western Finland HR, 1.18; 95% CI 1.02-1.36; unadjusted p = 0.03) with two additional taxa in the cluster, *C. symbiosum* and *C. glycyrrhizinilyticum*, which were not individually associated with type 2 diabetes in Western Finland data. In the beta diversity analysis, the first PC axis did not show an association with incident type 2 diabetes in the Western Finland data (HR, 0.94; 95% CI 0.79-1.11; unadjusted p = 0.45). Fewer participants in Western Finland with relative abundance of *C. citroniae* below Q1 developed incident type 2 diabetes than those above this quartile (**Figure 3**). For other associated features, a similar division was seen with an increased risk above the median (Q2) relative abundance.

**Figure 3.**
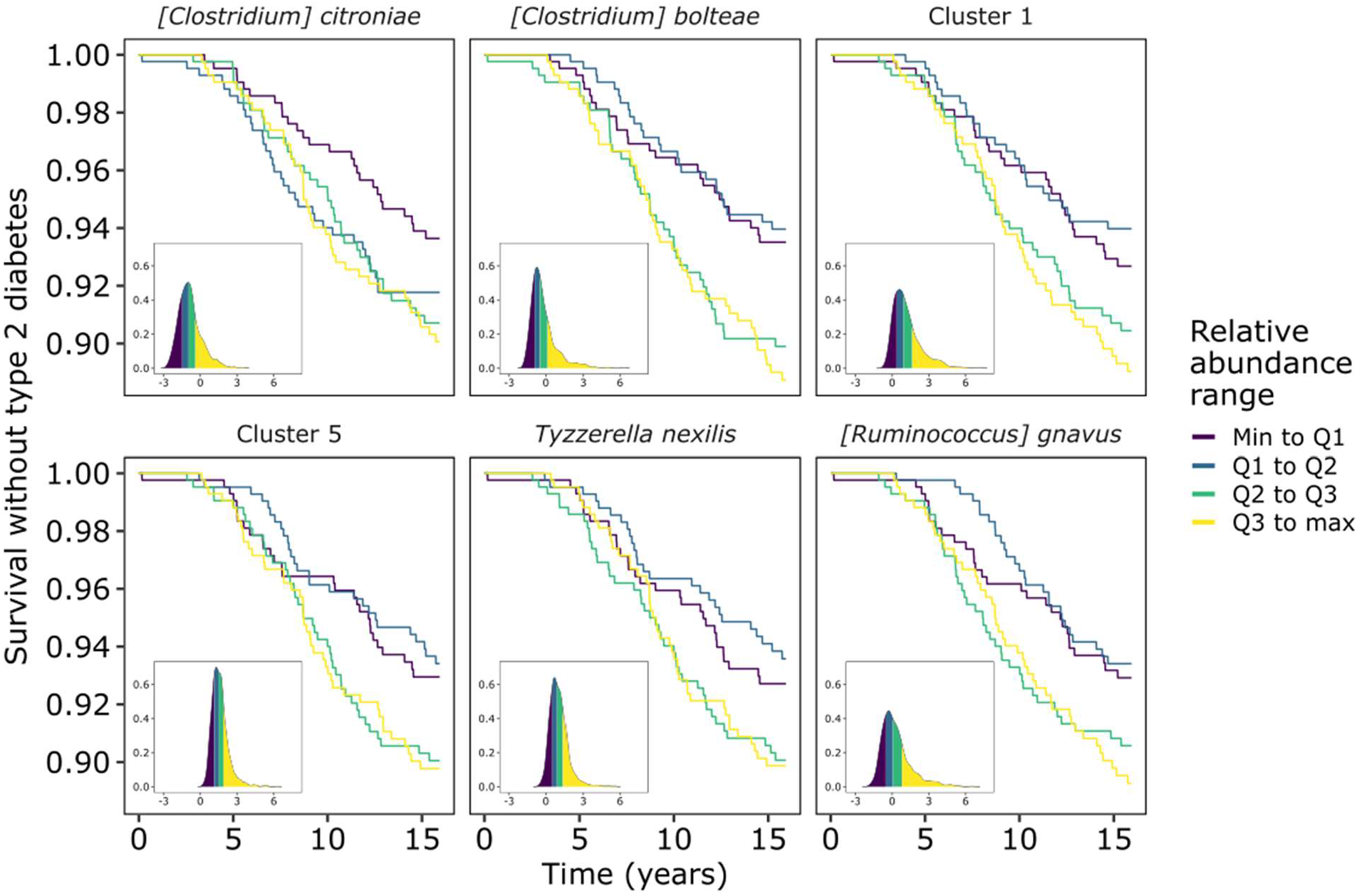
Kaplan-Meier curves for features with significant effect sizes in both data sets, displaying diabetes-free survival times of participants in Western Finland. Curves are separated by ranges between quartiles of relative abundance of each feature. Distribution of the participants with the same relative abundance ranges are included as inlays for each of the features.

## Conclusions

Previous studies have identified several biometric, genetic and lifestyle risk factors for incident type 2 diabetes and established their role in its development (25). Despite adjusting for several known risk factors, we demonstrated that several common taxa in the gut microbiome among healthy Finnish adults were associated with incident type 2 diabetes over a long-term follow-up. Specifically, we identified four species in the family *Lachnospiraceae* robustly associated with a higher type 2 diabetes risk in two geographically and genetically separate regions of Finland. Three of these taxa could be clustered together by proportional abundance in both geographic areas, and combined abundance of the 4 taxa was also predictive of incident type 2 diabetes.

Our findings are supported by several prior cross-sectional observations of microbiome composition related to type 2 diabetes and its risk factors. For example, *C. citroniae* has been positively associated with production of trimethylamine N-oxide (TMAO) production, likely connected with intake of red meat (26). The positive association between red meat intake and type 2 diabetes risk has been known for over 15 years (27). Furthermore, TMAO has been implicated in adipose tissue inflammation and impeded hepatic insulin signaling, which are connected with increased insulin resistance, high blood glucose levels, and type 2 diabetes (28). *C. bolteae* has been reported to be enriched in type 2 diabetes patients in previous cross-sectional study along with other opportunistic pathogens (4). Interestingly, the abundance of *C. bolteae* was reduced in patients treated with acarbose, an alpha-glucosidase inhibitor which is used as an antidiabetic drug (29). Acarbose works by inhibiting the breakdown of complex polysaccharides in the small intestine, which makes these compounds available for microbes in the colon and helps to lower blood glucose levels through the slower uptake of simple sugars. Also, the abundance of *T. nexilis* has been observed to decrease drastically in response to feeding participants a soluble fibre, polydextrose (30). Polydextrose supplementation in connection to high-fat diets has been reported to increase the concentration of postprandial plasma glucagon-like peptide-1, which is involved in regulation of blood glucose levels (31). Altogether, the abundance of *C. bolteae* and *T. nexilis* appears to be related to intake and availability of different polysaccharides in the colon, which likely influences their ecological niche. However, the mechanistic details of the link between these taxa and blood glucose levels remains to be clarified in detail. The abundance of *R*.*gnavus* is potentially related to glucose metabolism regulation and linked to increases in inflammatory cytokines, both of which are related to type 2 diabetes pathophysiology (5,32).

All four observed diabetes-associated taxa have been previously linked with other metabolic diseases and risk factors. For example, *R. gnavus* has been positively associated with obesity in animals (33,34), and humans (35). These taxa have also been associated with serum gamma-glutamyl transferase levels, an important liver disease marker (36). Our previous cross-sectional study of fatty liver disease in FINRISK 2002 also features serum gamma-glutamyl transferase level as a component of the modeled risk index and detected positive associations of all four taxa with a higher disease risk (37). Thus, the results of the current study support several links between dietary habits, metabolic diseases, and type 2 diabetes, likely modulated by the gut microbiome.

While only a subset of the associations with individual gut microbiome taxa in Eastern Finland were detected in Western Finland, remarkably, the 18 taxa associated with type 2 diabetes in the East clustered identically in the West (**Figure 1**). The association directions of the features with incident type 2 diabetes were mostly consistent between East and West data, also for features with statistically inconclusive results (**Figure 2**). However, there were also several taxa with inconsistent results between the two data.

Interestingly, for the robustly associated features the difference in type 2 diabetes incidence among the relative abundance quartiles emerges only after around 5 years of follow-up (**Figure 3**). Thus, it might have been challenging to detect taxa associations in previous studies with shorter follow-up times (7) or cross-sectional settings. Furthermore, the relative abundance distributions of all associated features are slightly positively skewed, with long tails of higher values. For features other than *C. citroniae*, the long-term risk of incident type 2 diabetes, however, seems to be increasing only after relative abundance values above the median. The relative abundance of *C. citroniae* is however quite low compared to the other taxa (or clusters), and only participants below Q1 seem to have a lower risk of developing incident type 2 diabetes. Thus, the metabolism of *C. citroniae*, such as its potential for TMAO production (26,38), might be important for the pathogenesis of type 2 diabetes.

The two species historically classified in the genus *Clostridium* (*C. citroniae* and *C. bolteae*) have recently been reclassified into a new genus, *Enterocloster* (39). This close phylogenetic relatedness might further indicate sharing of metabolic traits between these taxa. Also, other members of this new genus, such as *C. clostridioforme*, have been associated with metabolic diseases such as fatty liver disease (37), and with production of TMAO (26). Additionally, *C. clostridioforme* and *C. symbiosum* produce 3-methyl-4-(trimethylammonio)butanoate (3M-4-TMAB) and 4-(trimethylammonio)pentanoate (4-TMAP) which were reported to be mechanistically linked to type 2 diabetes pathogenesis (40). These connections could warrant further study on the members in the new genus *Enterocloster* and their connections with chronic diseases. Furthermore, many of the taxa associations in our study have only been previously observed with shotgun metagenomics (4,26,29,30,36). For example, to the best of our knowledge *C. bolteae* and *T. nexilis* have not been associated with type 2 diabetes in studies where 16S rRNA amplicon sequencing has been used. Studies reporting associations between type 2 diabetes and the gut microbiome composition should thus preferably utilize *e*.*g*., full-length 16S rRNA gene sequencing, or shotgun metagenomics, instead of 16S rRNA metabarcoding. For example, the construction of microbiome risk scores for type 2 diabetes is a promising approach to aid in its diagnosis and prevention (7). However, this method would also benefit from higher taxonomic resolution enabled by shotgun metagenomic or full length 16S rRNA gene sequencing instead of amplicon sequencing.

The strengths of the current study include the high taxonomic coverage and resolution of shotgun sequencing, long follow-up time, and a large, unselected study sample. In addition, the detected microbial signals support several previous cross-sectional observations on connections between the gut microbiome and type 2 diabetes and are robust in the geographically and genetically distinct regions in Finland. The results were also not confounded by anti-diabetic drugs, including metformin. Thus, these taxa are more likely to be associated with type 2 diabetes progression or onset, than related to the effects of dysglycemia (9). However, the use of shallow shotgun metagenomics also limits the current study to describing associations between taxa and incident disease, because the depth of the sequencing prevents genome assembly. We note that prospective studies with long follow-up times of over 5 years can be a powerful tool to detect early signals of diseases with known connections to gut microbiome composition.

We are not aware of previous long-term prospective studies on the associations between type 2 diabetes and the gut microbiome, similar to the current study. Thus, our results should be further validated with studies in suitable cohorts to address their generalizability. Further laboratory experiments with animal models could likely establish the required mechanistic and causal evidence to link specific microbial species and strains conclusively to type 2 diabetes pathogenesis. The current study thus serves as a stepping stone towards the goal of improved prediction and the development of effective treatments for type 2 diabetes through modification of the gut microbiome.

## Data Availability

The source code used to analyze the data and produce our results is included in the
Supplementary Information. Due to sensitive health information of individuals, the datasets
analyzed during the current study are not public, but are available based on a written application
to the THL Biobank as instructed in: https://thl.fi/en/web/thl-biobank/for-researchers

## Acknowledgements

M.O.R., P.P.E., V.S., R.K, L.L and T.J.N designed the work. A.S.H., G.M., P.J., V.S. and R.K. acquired the data. M.O.R., P.P.E., L.L. and T.J.N. analyzed the data. M.O.R. and P.P.E. wrote the manuscript in consultation with all authors. M.I., P.J., V.S., R.K., L.L. and T.J.N. supervised the work. All authors gave final approval of the version to be published. T.J.N. is the guarantor of this work and, as such, had full access to all the data in the study and takes responsibility for the integrity of the data and the accuracy of the data analysis. Furthermore, we would like to thank all participants of the FINRISK 2002 study, and Tara Schwartz for assistance with laboratory work.

This research was supported in part by grants from the Finnish Cultural Foundation, the Finnish Foundation for Cardiovascular Research, the Emil Aaltonen Foundation, the Finnish Medical Foundation, the Sigrid Juselius Foundation, and the Academy of Finland (#338818 to M.O.R.; #321356 to A.S.H.; #295741, #307127 to L.L.; #321351 to T.J.N.). Additional support was provided by Illumina, Inc. and Janssen Pharmaceutica through their sponsorship of the Center for Microbiome Innovation at UCSD.

The authors disclose the following potential conflicts-of-interest: T.J.N. has received speaking honoraria from Servier. V.S. has consulted for Sanofi and received honorarium from this company. He also has ongoing research collaboration with Bayer AG, all unrelated to this study.

## Notes

### Author Declarations

The Coordinating Ethics Committee of the Helsinki University Hospital District approved the study protocol for FINRISK 2002 (Ref. 558/E3/2001).

