## Supplementary material for "Gut microbiome composition is predictive of incident type 2 diabetes": Analysis_code_v1.0.R

```
#Analysis scripts for the manuscript: "Gut microbiome composition is predictive of incident type 2 diabetes" by Ruuskanen & Erawijantari et al.
#Due to sensitive health information, the data in this study are available based on a written application to the THL Biobank as instructed in: https://thl.fi/en/web/thl-biobank/for-researchers

if (!requireNamespace("BiocManager")) {
  install.packages("BiocManager")
}

#Use development version of ComplexHeatmap, 2.7.11<
#library(devtools)
#install_github("jokergoo/ComplexHeatmap")
#devtools::install_github("slowkow/ggrepel")

packages <- c("ggplot2", "biomformat", "ggthemes", "phyloseq", "vegan", "uwot", "patchwork", "microbiome", "tidyverse", "reshape2", "survival", "magrittr", "ggnewscale", "propr", "ComplexHeatmap", "maptree", "RColorBrewer", "rms", "viridis", "scales", "data.table")


is.installed <- function(pkg) {
  new.pkg <- pkg[!(pkg %in% installed.packages()[, "Package"])]
  if (length(new.pkg)) {
    BiocManager::install(new.pkg, ask=F)
  }
  sapply(pkg, require, character.only = TRUE)
}
is.installed(packages)

wideScreen <- function(howWide=Sys.getenv("COLUMNS")) {
  options(width=as.integer(howWide))
}
wideScreen()

theme_set(theme_tufte(base_family = "sans", base_size = 18) + theme(panel.border = element_rect(colour = "black", fill = NA), axis.text = element_text(colour = "black", size = 18)))


#All data are included in the THL Biobank release package.
#Phenotype data is loaded from the included R object
load("FR_02_phenotype_data.RData")
#Subset to data which includes the fecal samples
FR02 <- FR02[!is.na(FR02$Barcode),]
row.names(FR02) <- FR02$Barcode
#Construct objects with NCBI data from SHOGUN
if (file.exists("microbiome_predicts_incident_T2D/ncbi_data.RDs")) {
  ncbi_data <- readRDS("microbiome_predicts_incident_T2D/ncbi_data.RDs")
} else {
  #Construct the primary phyloseq object and subset to FR02 samples.
  ncbi_data <- biomformat::read_biom("microbiome_predicts_incident_T2D/combined_redist.species.biom") #BIOM table from the SHOGUN species-level output
  ncbi_data <- biomformat::biom_data(ncbi_data)
  ncbi_tax_table <- strsplit(row.names(as.matrix(ncbi_data)), ";")
  ncbi_tax_table <- matrix(unlist(ncbi_tax_table), nrow=length(ncbi_tax_table), byrow=T)
  row.names(ncbi_data) <- row.names(ncbi_tax_table)
  ncbi_data <- phyloseq(otu_table(as.matrix(ncbi_data), taxa_are_rows=T), tax_table(ncbi_tax_table))
  #Format the tax table.
  colnames(tax_table(ncbi_data)) <- c("Domain", "Phylum", "Class", "Order", "Family",  "Genus", "Species")
  #Combine the phenotype data with the taxa data
  ncbi_data <- phyloseq(otu_table(ncbi_data), tax_table(ncbi_data), sample_data(FR02))
  #remove samples with less than 50k reads (total)
  to_be_pruned <- sample_sums(ncbi_data) > 50000
  ncbi_data <- prune_samples(to_be_pruned, ncbi_data)
  #remove pregnant (GRAVID==2) participants
  ncbi_data <- subset_samples(ncbi_data, GRAVID %in% c(1, NA))
  #remove participants who have used antibiotics in the last 6 months (BL_USE_RX_J01==1)
  ncbi_data <- subset_samples(ncbi_data, BL_USE_RX_J01 %in% c(0, NA))
  #remove participants with prevalent diabetes (PREVAL_DIAB==1)
  ncbi_data <- subset_samples(ncbi_data, PREVAL_DIAB==0)
  #remove participants with diabetes indicator values over set guidelines: FR02_GLUK_NOLLA >= 7, FR02_GLUK_120 >= 11.1 & HBA1C >= 48 (ignore NA values)
  ncbi_data <- subset_samples(ncbi_data, FR02_GLUK_NOLLA<7 | is.na(FR02_GLUK_NOLLA))
  ncbi_data <- subset_samples(ncbi_data, FR02_GLUK_120<11.1 | is.na(FR02_GLUK_120))
  ncbi_data <- subset_samples(ncbi_data, HBA1C<48 | is.na(HBA1C))
  #save the final objects
  saveRDS(ncbi_data, "microbiome_predicts_incident_T2D/ncbi_data.RDs")
}


#Functions
prediab_cat <- function(pseq){
	data <- sample_data(pseq)
	prediab <- ifelse(data$FR02_GLUK_NOLLA >= 5.6 & data$FR02_GLUK_NOLLA < 6.9 | data$FR02_GLUK_120 >= 7.8 & data$FR02_GLUK_120 < 11 | data$HBA1C >= 39 & data$HBA1C < 47, 1, 0)
	prediab <- as.factor(prediab)
    return(prediab)
}

cox_wrapper <- function(data,
                        predictors,
                        covariates,
                        status,
                        time_to_event,
                        alpha_level,
                        normalize,
                        test_ph_assumption) {
  if(normalize) {  
    if(class(data[, predictors]) == "numeric") {
      x <- data[, predictors]
      data[, predictors] <- (x - mean(x, na.rm = T))/sd(x, na.rm = T) 
    } else {
      data[, predictors] <- apply(data[, predictors], 2, FUN = function(x) {(x - mean(x, na.rm = T))/sd(x, na.rm = T) })
    }
  }
  ## Formulas ***************************
  linear_formulas <- lapply(predictors, function(x) {
    formula_data <- deparse(substitute(data))
    formula <- paste0("Surv(", formula_data, "$", time_to_event, ", ",formula_data,"$", status, ") ~ ",paste(covariates, collapse = "+"), " + ",x)
    return(formula)
  }) %>% 
    set_names(predictors)
  ## Cox regression *********************
  print("Cox")
  linear_cox_fit <- lapply(linear_formulas, function(x) {
    coxph(as.formula(x), data=data, x=TRUE)
  })
  ## Check PH assumptions ****************
  if(test_ph_assumption) {
    print("PH assumptions")
    ph_assumption <-  lapply(predictors, function(m) {
      west <- cox.zph(linear_cox_fit[[m]])
      p_values <- west$table[, "p"]
      # significant cases
      x <- which(p_values < 1)
      if(length(x) == 0) {
        return(NULL)
      }
      df <- data.frame(feature = m, variable_not_ph = names(x), p_value = p_values[x])
    }) %>% 
      do.call(rbind, .) %>%
      mutate(p_adj = p.adjust(p_value, "BH")) %>% 
      filter(p_value < alpha_level)
  }
  ## Results *****************************
  print("Results")
  results <- lapply(predictors, function(x) {
    df <- summary(linear_cox_fit[[x]])$coefficients %>% as.data.frame()
    df <- df[nrow(df), ] %>% 
      select(coef, "se(coef)", "z", "Pr(>|z|)") %>% 
      set_colnames(c("coef", "se_coef", "west_stat_value", "p")) %>% 
      mutate(west_stat = "Wald")
    df <- df %>% 
      mutate(predictor = x)
  }) %>% 
    do.call(rbind, .) 
  # Multiple westing correction
  results <- results %>%
    mutate(P_adjusted = p.adjust(p, "BH")) %>% 
    ungroup() %>% 
    group_by(predictor) 
  # Results in neat form for presentation
  neat_results <- results %>% 
    # filter(p == min(p)) %>% 
    ungroup() %>% 
    mutate(HR = round(exp(coef),3)) %>% 
    mutate(HR_lower_95 = round(exp(coef - 1.96*se_coef), 3),
           HR_upper_95 = round(exp(coef + 1.96*se_coef), 3),
           P = round(p, 5),
           Coefficient = round(coef, 3),
           "Coefficient SE" = round(se_coef, 3)) %>% 
    mutate(HR = paste0(HR, " (95% CI, ", HR_lower_95, "-", HR_upper_95, ")")) %>% 
    select(Predictor = predictor, Coefficient, "Coefficient SE", HR, "p","P_adjusted", "west_stat_value", "west_stat") %>%
    mutate(HR = ifelse(is.na(Coefficient), NA, HR))  %>% 
    filter(P_adjusted < alpha_level) %>% 
    arrange(P_adjusted) %>% 
    set_colnames(c("Predictor", "Coefficient", "Coefficient SE", "HR","P-value" ,"P (adjusted)", "west Statistic Value", "west Statistic"))
  # Results in a form more convenient for further manipulations
  results <- results %>% 
    ungroup %>% 
    mutate(PH = exp(coef)) %>% 
    mutate(p_adj = P_adjusted) %>% 
    mutate(direction = ifelse(coef < 0, "negative", "positive"))
  if(nrow(neat_results) == 0) {
    return(list(results = results))
  }
  if(test_ph_assumption) {
    if(nrow(neat_results) == 0) {
      return(list(results = results, ph_assumption = ph_assumption))
    }
    return(list(neat_results = neat_results, 
                results = results,
                ph_assumption = ph_assumption))
  }
  return(list(neat_results = neat_results, results = results))
}

#preprocess data
if (file.exists("microbiome_predicts_incident_T2D/ncbi_data_raw_east.RDs") && file.exists("microbiome_predicts_incident_T2D/ncbi_data_raw_west.RDs") &&
file.exists("microbiome_predicts_incident_T2D/ncbi_data_main.RDs") && file.exists("microbiome_predicts_incident_T2D/ncbi_pca.RDs") && 
file.exists("microbiome_predicts_incident_T2D/ncbi_data_east.RDs") && file.exists("microbiome_predicts_incident_T2D/ncbi_pca_east.RDs") &&
file.exists("microbiome_predicts_incident_T2D/ncbi_data_west.RDs") && file.exists("microbiome_predicts_incident_T2D/ncbi_pca_west.RDs") &&
file.exists("microbiome_predicts_incident_T2D/ncbi_pca_data_east.RDs") && file.exists("microbiome_predicts_incident_T2D/ncbi_pca_data_west.RDs")) {
  ncbi_data_raw_east <- readRDS("microbiome_predicts_incident_T2D/ncbi_data_raw_east.RDs")
  ncbi_data_raw_west <- readRDS("microbiome_predicts_incident_T2D/ncbi_data_raw_west.RDs")
  ncbi_data_main <- readRDS("microbiome_predicts_incident_T2D/ncbi_data_main.RDs")
  ncbi_data_east <- readRDS("microbiome_predicts_incident_T2D/ncbi_data_east.RDs")
  ncbi_data_west <- readRDS("microbiome_predicts_incident_T2D/ncbi_data_west.RDs")
  ncbi_pca <- readRDS("microbiome_predicts_incident_T2D/ncbi_pca.RDs")
  ncbi_pca_east <- readRDS("microbiome_predicts_incident_T2D/ncbi_pca_east.RDs")
  ncbi_pca_west <- readRDS("microbiome_predicts_incident_T2D/ncbi_pca_west.RDs")
  ncbi_pca_data_east <- readRDS("microbiome_predicts_incident_T2D/ncbi_pca_data_east.RDs")
  ncbi_pca_data_west <- readRDS("microbiome_predicts_incident_T2D/ncbi_pca_data_west.RDs")
} else {
  #Limit taxa to core in the east (EAST) set
  core_ncbi_taxa <- core(prune_samples(meta(ncbi_data)$EAST == 1, ncbi_data) %>% transform("compositional"), detection = .1/100, prevalence = 1/10) %>% taxa_names()
  ncbi_data_main <- prune_taxa(core_ncbi_taxa, ncbi_data)
  #divide non-transformed data to east/west (EAST/WEST) sets
  ncbi_data_raw_east <- prune_samples(meta(ncbi_data_main)$EAST == 1, ncbi_data_main)
  ncbi_data_raw_west <- prune_samples(meta(ncbi_data_main)$EAST == 0, ncbi_data_main)
  #CLR-transform raw counts
  ncbi_data_main <- transform(ncbi_data_main, "clr")
  #calculate additional variables
  PREDIAB <- prediab_cat(ncbi_data_main)
  NON_HDL <- sample_data(ncbi_data)$KOL - sample_data(ncbi_data)$HDL
  #calculate diversity
  ncbi_diversity <- estimate_richness(ncbi_data, measures = c("Observed", "Shannon"))
  #reduce metadata to useful columns
  useful_variables <- c("BL_AGE", "BMI", "MEN", "SYSTM", "CURR_SMOKE", "TRIG", "INCIDENT_DIAB_T2", "DIAB_T2_AGEDIFF", "EAST")
  sample_data(ncbi_data_main) <- sample_data(ncbi_data_main)[,sample_variables(ncbi_data_main) %in% useful_variables]
  #separate transformed and curated data to east/west (EAST/WEST) sets 
  ncbi_data_east <- prune_samples(meta(ncbi_data_main)$EAST == 1, ncbi_data_main)
  ncbi_data_west <- prune_samples(meta(ncbi_data_main)$EAST == 0, ncbi_data_main)
  #calculate 10 first PCAs with full community
  ncbi_data_raw_clr <- transform(ncbi_data, "clr")
  ncbi_pca <- ordinate(ncbi_data_raw_clr, "RDA")
  ncbi_pca_data <- as.data.frame(scores(ncbi_pca, choices = c(1:10))$sites)
  ncbi_pca_east <- ordinate(prune_samples(meta(ncbi_data_raw_clr)$EAST == 1, ncbi_data_raw_clr), "RDA")
  ncbi_pca_data_east <- as.data.frame(scores(ncbi_pca_east, choices = c(1:10))$sites)
  ncbi_pca_west <- ordinate(prune_samples(meta(ncbi_data_raw_clr)$EAST == 0, ncbi_data_raw_clr), "RDA")
  ncbi_pca_data_west <- as.data.frame(scores(ncbi_pca_west, choices = c(1:10))$sites)
  #combine with additional data
  sample_data(ncbi_data_main) <- cbind(sample_data(ncbi_data_main), PREDIAB, NON_HDL, ncbi_diversity, ncbi_pca_data)
  sample_data(ncbi_data_east) <- cbind(sample_data(ncbi_data_east), PREDIAB = PREDIAB[which(meta(ncbi_data_main)$EAST == 1)], NON_HDL = NON_HDL[which(meta(ncbi_data_main)$EAST == 1)], ncbi_diversity[which(meta(ncbi_data_main)$EAST == 1),], ncbi_pca_data_east)
  sample_data(ncbi_data_west) <- cbind(sample_data(ncbi_data_west), PREDIAB = PREDIAB[which(meta(ncbi_data_main)$EAST == 0)], NON_HDL = NON_HDL[which(meta(ncbi_data_main)$EAST == 0)], ncbi_diversity[which(meta(ncbi_data_main)$EAST == 0),], ncbi_pca_data_west)
  saveRDS(ncbi_data_raw_east, "microbiome_predicts_incident_T2D/ncbi_data_raw_east.RDs")
  saveRDS(ncbi_data_raw_west, "microbiome_predicts_incident_T2D/ncbi_data_raw_west.RDs")
  saveRDS(ncbi_data_main, "microbiome_predicts_incident_T2D/ncbi_data_main.RDs")
  saveRDS(ncbi_data_east, "microbiome_predicts_incident_T2D/ncbi_data_east.RDs")
  saveRDS(ncbi_data_west, "microbiome_predicts_incident_T2D/ncbi_data_west.RDs")
  saveRDS(ncbi_pca, "microbiome_predicts_incident_T2D/ncbi_pca.RDs")
  saveRDS(ncbi_pca_east, "microbiome_predicts_incident_T2D/ncbi_pca_east.RDs")
  saveRDS(ncbi_pca_west, "microbiome_predicts_incident_T2D/ncbi_pca_west.RDs")
  saveRDS(ncbi_pca_data_east, "microbiome_predicts_incident_T2D/ncbi_pca_data_east.RDs")
  saveRDS(ncbi_pca_data_west, "microbiome_predicts_incident_T2D/ncbi_pca_data_west.RDs")
}

#Filter features based on corrected p-values in the east dataset
#set variables
alpha_level <- 0.05 #to filter
status <- "INCIDENT_DIAB_T2"
time_to_event <- "DIAB_T2_AGEDIFF"
ncbi_cox_data_east <- cbind(meta(ncbi_data_east), as.matrix(t(otu_table(ncbi_data_east))))
predictors <- c("Shannon", "Observed", colnames(ncbi_pca_data_east), taxa_names(ncbi_data_east))
covariates <- c("BL_AGE", "BMI", "MEN", "SYSTM", "NON_HDL", "CURR_SMOKE", "TRIG")
splines <- TRUE
normalize <- TRUE
test_ph_assumption <- FALSE
#Cox regression with previously defined function
set.seed(11235)
ncbi_cox_east <- cox_wrapper(data = ncbi_cox_data_east,
                        predictors = predictors,
                        covariates = covariates,
                        alpha_level = alpha_level,
                        status = status,
                        time_to_event = time_to_event,
                        normalize = normalize,
                        test_ph_assumption = test_ph_assumption)

ncbi_cox_results_east <- merge(ncbi_cox_east$neat_results, as.data.frame(ncbi_data_east@tax_table@.Data), by.x="Predictor", by.y="row.names")
ncbi_cox_results_east <- ncbi_cox_results_east[order(-ncbi_cox_results_east$Coefficient),]
ncbi_cox_results_east$Species <- gsub("s__", "", ncbi_cox_results_east$Species)
ncbi_cox_results_east$Species <- gsub("_", " ", ncbi_cox_results_east$Species)
ncbi_cox_results_east$Family <- gsub("f__", "", ncbi_cox_results_east$Family)

#Correlations and clustering between the associated taxa in east data
otu_table_assoc_taxa <- as.data.frame(otu_table(prune_taxa(ncbi_cox_east$neat_results$Predictor, ncbi_data_raw_east)))
rownames(otu_table_assoc_taxa) <- ncbi_cox_results_east$Species[match(rownames(otu_table_assoc_taxa), ncbi_cox_results_east$Predictor)]
set.seed(11235)
proprmatrix <- propr(t(otu_table_assoc_taxa), metric = "rho", p = 100)
clusters_assoc <- hclust(dist(proprmatrix@matrix), method = "ward.D2")
#Compute the Kelley-Gardner-Sutcliffe penalty function for a hierarchical cluster tree, to determine optimal number of clusters
op_k <- kgs(clusters_assoc, dist(proprmatrix@matrix), maxclus = 20)
op_k <- as.numeric(names(op_k[which(op_k == min(op_k))]))
cluster_ids <- cutree(tree = clusters_assoc, k = op_k)
svg("microbiome_predicts_incident_T2D/clusters.svg", width=10, height=10)
plot(clusters_assoc)
rect.hclust(clusters_assoc, k = op_k, border = 2:7)
dev.off()

heatmap_annotation <- data.frame(Species = rownames(proprmatrix@matrix), Cluster = cluster_ids)
heatmap_annotation$Predictor <- ncbi_cox_results_east$Predictor[match(heatmap_annotation$Species, ncbi_cox_results_east$Species)]

#Clustering correlating significant taxa for east and west data
#Combine read counts of clusters and calculate their CLR values
taxa_clusters <- merge(heatmap_annotation[c("Cluster")], ncbi_cox_results_east[c("Species", "Predictor")], by.x = "row.names", by.y = "Species")
taxa_clusters$Cluster <- factor(taxa_clusters$Cluster, levels = 1:length(unique(taxa_clusters$Cluster)))

cluster_phylo_east <- ncbi_data_raw_east
cluster_phylo_west <- ncbi_data_raw_west
index_taxa <- c()
for (cluster in levels(taxa_clusters$Cluster)) {
taxa_to_merge <- taxa_clusters$Predictor[which(taxa_clusters$Cluster == cluster)]
cluster_phylo_east <- merge_taxa(cluster_phylo_east, taxa_to_merge, archetype=1)
cluster_phylo_west <- merge_taxa(cluster_phylo_west, taxa_to_merge, archetype=1)
index_taxa[cluster] <- taxa_to_merge[1]
}
cluster_phylo_east <- transform(cluster_phylo_east, "clr")
cluster_phylo_west <- transform(cluster_phylo_west, "clr")
#Retain only clusters
cluster_phylo_east <- prune_taxa(index_taxa, cluster_phylo_east)
cluster_phylo_west <- prune_taxa(index_taxa, cluster_phylo_west)
taxa_names(cluster_phylo_east) <- paste0("Cluster_", taxa_clusters$Cluster[match(taxa_names(cluster_phylo_east), taxa_clusters$Predictor)])
taxa_names(cluster_phylo_west) <- paste0("Cluster_", taxa_clusters$Cluster[match(taxa_names(cluster_phylo_west), taxa_clusters$Predictor)])

#test the individual taxa and clusters in the east data
#set variables
alpha_level <- 1 #to include everything in the results
status <- "INCIDENT_DIAB_T2"
time_to_event <- "DIAB_T2_AGEDIFF"
ncbi_cox_data_east_2 <- cbind(meta(ncbi_data_east), as.matrix(t(otu_table(ncbi_data_east))), as.matrix(t(otu_table(cluster_phylo_east))))
predictors <- c(ncbi_cox_results_east$Predictor, taxa_names(cluster_phylo_east), "PC1")
covariates <- c("BL_AGE", "BMI", "MEN", "SYSTM", "NON_HDL", "CURR_SMOKE", "TRIG")
splines <- TRUE
normalize <- TRUE
test_ph_assumption <- FALSE
#Cox regression with previously defined function
set.seed(11235)
ncbi_cox_east_2 <- cox_wrapper(data = ncbi_cox_data_east_2,
                        predictors = predictors,
                        covariates = covariates,
                        alpha_level = alpha_level,
                        status = status,
                        time_to_event = time_to_event,
                        normalize = normalize,
                        test_ph_assumption = test_ph_assumption)

ncbi_cox_results_east_2 <- data.frame(ncbi_cox_east_2$neat_results)
ncbi_cox_results_east_2 <- merge(ncbi_cox_results_east_2[c("Predictor", "Coefficient", "HR", "P.value")], ncbi_cox_results_east[c("Predictor", "Family", "Species")], by = "Predictor", all = TRUE)
ncbi_cox_results_east_2 <- ncbi_cox_results_east_2[order(-ncbi_cox_results_east_2$Coefficient),]
ncbi_cox_results_east_2$Set <- "East"

#test the individual taxa and clusters in the west data
#use same variables as for previous model run (thus not repeated here)
ncbi_cox_data_west <- cbind(meta(ncbi_data_west), as.matrix(t(otu_table(ncbi_data_west))), as.matrix(t(otu_table(cluster_phylo_west))))
#Cox regression with previously defined function
set.seed(11235)
ncbi_cox_west <- cox_wrapper(data = ncbi_cox_data_west,
                        predictors = predictors,
                        covariates = covariates,
                        alpha_level = alpha_level,
                        status = status,
                        time_to_event = time_to_event,
                        normalize = normalize,
                        test_ph_assumption = test_ph_assumption)

ncbi_cox_results_west <- data.frame(ncbi_cox_west$neat_results)
ncbi_cox_results_west <- merge(ncbi_cox_results_west[c("Predictor", "Coefficient", "HR", "P.value")], ncbi_cox_results_east[c("Predictor", "Family", "Species")], by = "Predictor", all = TRUE)
ncbi_cox_results_west <- ncbi_cox_results_west[order(-ncbi_cox_results_west$Coefficient),]
ncbi_cox_results_west$Set <- "West"

#save results
results_out_east <- rbind(data.frame(ncbi_cox_east$neat_results), data.frame(ncbi_cox_east_2$neat_results[which(grepl("Cluster", ncbi_cox_east_2$neat_results$Predictor)),]))
results_out_west <- data.frame(ncbi_cox_west$neat_results)
results_out_east <- merge(results_out_east[c("Predictor", "Coefficient", "HR", "P.value", "P..adjusted.")], as.data.frame(ncbi_data_east@tax_table@.Data)["Species"], by.x="Predictor", by.y="row.names", all.x = TRUE)
results_out_west <- merge(results_out_west[c("Predictor", "Coefficient", "HR", "P.value", "P..adjusted.")], as.data.frame(ncbi_data_west@tax_table@.Data)["Species"], by.x="Predictor", by.y="row.names", all.x = TRUE)
results_out_west$P..adjusted. <- NA
results_out_east[which(grepl("Cluster", results_out_east$Predictor)),]$P..adjusted. <- NA
results_out <- merge(results_out_east, results_out_west, by="Predictor", suffixes=c(".east",".west"))
result_order <- results_out[rev(order(results_out$Coefficient.east)),]$Predictor
result_order <- c("PC1", paste0("Cluster_", 1:5),result_order[which(grepl("sp", result_order))])
results_out <- results_out[match(result_order, results_out$Predictor),]
results_out[-which(is.na(results_out$Species.east)),"Predictor"] <- as.character(results_out[-which(is.na(results_out$Species.east)),"Species.east"])
results_out$Predictor <- gsub("s__", "", results_out$Predictor)
results_out$Predictor <- gsub("_", " ", results_out$Predictor)
names(results_out) <- gsub("\\.\\.", ".", names(results_out))
results_out <- results_out[,!names(results_out) %in% c("Species.east", "Species.west", "P.adjusted.west")]
results_out[c("P.value.east", "P.adjusted.east", "P.value.west")] <- lapply(results_out[c("P.value.east", "P.adjusted.east", "P.value.west")], function (x) round(x, 4))
write.csv(results_out, "microbiome_predicts_incident_T2D/Table_S1.csv", row.names=F)

#plot heatmap of taxa associations, clustering, and hazard ratios in the east data
newnames <- lapply(rownames(proprmatrix@matrix),function(x) bquote(italic(.(x))))
heatmap_annotation$HR <- gsub("([0-9]\\.[0-9]*)[[:space:]].*", "\\1", ncbi_cox_results_east$HR[match(heatmap_annotation$Predictor, ncbi_cox_results_east$Predictor)])
heatmap_annotation$HR <- round(as.numeric(as.character(heatmap_annotation$HR)), 1)
heatmap_annotation$HR <- factor(heatmap_annotation$HR, levels = rev(seq(0.8, 1.2, 0.1)))

ann_colors <- list(HR = brewer.pal(n = 5, name = "BrBG"), Cluster = brewer.pal(n = 10, name = "Paired")[-seq(1,9,2)])
names(ann_colors$HR) <- levels(heatmap_annotation$HR)
names(ann_colors$Cluster) <- c("1", "2", "3", "4", "5")
ann_colors$Cluster <- factor(ann_colors$Cluster, levels = ann_colors$Cluster[c(4,2,5,6,3,1)])
heatmap_colors <- rev(brewer.pal(n = 10, name = "RdBu"))
heatmap_colors[c(5,6)] <- "#FFFFFF"
svg("microbiome_predicts_incident_T2D/correlations.svg", width=15, height=15)
pheatmap(proprmatrix@matrix, labels_row = as.expression(newnames), labels_col = as.expression(newnames), annotation_row = heatmap_annotation[4], treeheight_row = 0, annotation_col = heatmap_annotation[2], annotation_colors = ann_colors, cutree_rows = op_k, cutree_cols = op_k, clustering_method = "ward.D2", color = heatmap_colors, breaks = seq(-1, 1, length.out = 11), legend_breaks = seq(-1, 1, length.out = 11), cellwidth=10, cellheight=10)
dev.off()

#plot HR of both west and east data
ncbi_cox_results <- rbind(ncbi_cox_results_east_2, ncbi_cox_results_west)

Species <- c()
Family <- c()
Set <- c()
Facet <- c()
HR <- c()
HR1 <- c()
HR2 <- c()

for (i in 1:length(ncbi_cox_results$Predictor)){
  Species[[i]] <- ifelse(is.na(ncbi_cox_results$Species[i]), sub("_", " ", ncbi_cox_results$Predictor[i]), as.character(ncbi_cox_results$Species[i]))
  Family[[i]] <- ifelse(is.na(ncbi_cox_results$Family[i]), NA, as.character(ncbi_cox_results$Family[i]))
  HR[[i]] <- str_split(ncbi_cox_results$HR[i]," ")[[1]][1]
  HR_range <- str_split(ncbi_cox_results$HR[i]," ")[[1]][4]
  HR1[[i]] <- str_split(HR_range,"-")[[1]][1]
  HR2_bef <- str_split(HR_range,"-")[[1]][2]
  HR2[[i]] <- substr(HR2_bef,1,nchar(HR2_bef)-1)
  Set[[i]] <- ncbi_cox_results$Set[i]
  Facet[[i]] <- ifelse(is.na(ncbi_cox_results$Family[i]), "Grouping", "Taxa")
  HRdf <- data.frame(Species = Species,
					  Family = Family,
					  Set = Set,
					  Facet = Facet,
                      HR = HR,
                      HR1 = HR1,
                      HR2 = HR2)
}

family_color_map <- data.frame(Color = c("#7b562e", "#9bb940", "#c5bb9a", "darkred", "#ff4ae3", "#339a00", "#d78343", "#5f96d6", "black"),
 Family = c("Bacteroidaceae", "Clostridiaceae", "Eggerthellaceae", "Eubacteriaceae", "Lachnospiraceae", "Oscillospiraceae", "Rickenellaceae", "Sutterellaceae", NA))

HRdf$Species <- factor(HRdf$Species, levels = c(paste0("Cluster ", 5:1), "PC1", as.character(HRdf[which(HRdf$Set %in% "East" & HRdf$Facet %in% "Taxa"),][order(HRdf[which(HRdf$Set %in% "East" & HRdf$Facet %in% "Taxa"),]$HR),]$Species))) #order features by effect size in the east data
p <-  ggplot(data = HRdf, aes(y = Species, x = as.numeric(as.character(HR)), color = Family)) + 
      geom_pointrange(aes(xmin=as.numeric(as.character(HR1)), xmax=as.numeric(as.character(HR2))), lwd = 1) +
      scale_x_continuous(limits = c(0.6, 1.45)) +
	  scale_color_manual(name = "Family", values = as.character(family_color_map$Color)) +
	  guides(color = guide_legend(override.aes = list(size = 1.4))) +
      xlab("HR") + ylab("Species") +
      geom_vline(xintercept=c(1.0), linetype="dotted") +
	  theme(axis.text.y = element_text(face = "italic"), legend.text = element_text(face = "italic"), axis.title.y = element_blank()) +
	  facet_grid(Facet~Set, scales = "free")

ggsave("microbiome_predicts_incident_T2D/HR_comparison.svg", plot=p, units="in", width=15, height=10)

#Plot Kaplan-Meier curves
kp_predictors <- ncbi_cox_results_west$Predictor[which(ncbi_cox_results_west$P.value < 0.05)]
kp_covariates <- covariates
kp_time_to_event <- time_to_event
kp_status <- status
kp_data <- ncbi_cox_data_west[,which(colnames(ncbi_cox_data_west) %in% c(kp_status, kp_time_to_event, kp_predictors, kp_covariates))]
kp_time <- seq(0, max(kp_data$DIAB_T2_AGEDIFF), by = .01)
kp_list <- list(NULL)
for (time in 1:length(kp_time)) {
kp_table <- lapply(kp_predictors, function(x) {
    return_table <- data.frame(groupkm(kp_data[x], Surv(kp_data$DIAB_T2_AGEDIFF, kp_data$INCIDENT_DIAB_T2), g=4, u=kp_time[time], pl=FALSE))
	return_table$Predictor <- x
	return_table$quantile <- c(1:4)
    return(return_table)
  })
kp_table <- do.call(rbind, kp_table)
kp_table$time <- kp_time[time]
kp_list[[time]] <- kp_table
}

kp_list <- do.call(rbind, kp_list)
kp_predictors <- recode(kp_predictors, 'sp2673' = "[Clostridium] citroniae", 'sp2671' = "[Clostridium] bolteae", 'sp2697' = "Tyzzerella nexilis", 'sp2638' = "[Ruminococcus] gnavus")
kp_predictors <- gsub("_", " ", kp_predictors)
kp_list$Predictor <- recode(kp_list$Predictor, 'sp2673' = "[Clostridium] citroniae", 'sp2671' = "[Clostridium] bolteae", 'sp2697' = "Tyzzerella nexilis", 'sp2638' = "[Ruminococcus] gnavus")
kp_list$Predictor <- gsub("_", " ", kp_list$Predictor)
kp_list$Predictor <- factor(kp_list$Predictor, levels = kp_predictors)

p <-  ggplot(data = kp_list, aes(y = KM, x = time, group = quantile)) + 
	  geom_line(aes(color = quantile)) +
	  scale_color_viridis(labels = c("Min to Q1", "Q1 to Q2", "Q2 to Q3", "Q3 to max")) +
	  scale_y_continuous(breaks = pretty_breaks()) +
	  guides(color = guide_legend(override.aes = list(size = 1.4)), fill = "none") +
      xlab("Time (years)") + ylab("Survival without type 2 diabetes") +
	  labs(color = "Relative\nabundance\nrange") +
	  facet_wrap(~ Predictor)
ggsave("microbiome_predicts_incident_T2D/KP_plot.svg", plot=p, units="cm", width=30, height=20)

#Plot distributions of the quartiles (for inlays in the KP-plot)
quartile_data <- lapply(kp_data[ncbi_cox_results_west$Predictor[which(ncbi_cox_results_west$P.value < 0.05)]], 
	function(x) data.frame(x_value = density(x)$x, 
							y_value = density(x)$y,
							quartile = factor(paste0("Q",findInterval(density(x)$x,quantile(x, prob=c(0, 0.25, 0.5, 0.75, 1)), all.inside=T)))))
quartile_data <- data.frame(rbindlist(quartile_data, idcol="Predictor"))
quartile_data$Predictor <- recode(quartile_data$Predictor, 'sp2673' = "[Clostridium] citroniae", 'sp2671' = "[Clostridium] bolteae", 'sp2697' = "Tyzzerella nexilis", 'sp2638' = "[Ruminococcus] gnavus")
quartile_data$Predictor <- gsub("_", " ", quartile_data$Predictor)
quartile_data$Predictor <- factor(quartile_data$Predictor, levels = kp_predictors)

p <- ggplot(quartile_data, aes(x_value, y_value)) + 
	  geom_line() + 
	  geom_ribbon(aes(ymin=0, ymax=y_value, fill=quartile)) + 
	  scale_fill_viridis(labels = c("Q1", "Q2", "Q3", "Q4"), discrete=T) +
	  guides(fill = "none") +
	  theme(axis.title = element_blank()) +
	  facet_wrap(~ Predictor)
ggsave("microbiome_predicts_incident_T2D/KP_plot_quartiles.svg", plot=p, units="cm", width=30, height=20)

#Format and print Table 1
table_data <- data.table(meta(ncbi_data))
table_data$NON_HDL <- table_data$KOL - table_data$HDL
table_variables <- c("BL_AGE", "BMI", "SYSTM", "NON_HDL", "FR02_GLUK_NOLLA", "FR02_GLUK_120", "HBA1C", "TRIG")
table_variables2 <- c("CURR_SMOKE", "MEN", "EAST")
table_data <- table_data[, .SD, .SDcols = c(table_variables, table_variables2, "INCIDENT_DIAB_T2")]
table_data$MEN <- ifelse(table_data$MEN == 0, 1, 0) #invert the variable to count female (not male) participants as 1's for the table
table1 <- transpose(merge(rbind(table_data[, c(INCIDENT_DIAB_T2 = "all", lapply(.SD, function(x) paste0(round(mean(x, na.rm=T),1), "+-", round(sd(x, na.rm=T),1))) ), .SDcols=table_variables],
	table_data[, lapply(.SD, function(x) paste0(round(mean(x, na.rm=T),1), "+-", round(sd(x, na.rm=T),1))), by=INCIDENT_DIAB_T2, .SDcols=table_variables]),
	rbind(table_data[, c(INCIDENT_DIAB_T2 = "all", lapply(.SD, function(x) paste0(table(x)[2], " (", round(table(x)[2]/length(x)*100, 1), ")")) ), .SDcols=table_variables2],
	table_data[, lapply(.SD, function(x) paste0(table(x)[2], " (", round(table(x)[2]/length(x)*100, 1), ")")), by=INCIDENT_DIAB_T2, .SDcols=table_variables2]), by = "INCIDENT_DIAB_T2"), keep.names = "col", make.names = "INCIDENT_DIAB_T2")
table1 <- table1[,c("col", "all", "1", "0")]
p_values_cont <- as.vector(as.matrix(table_data[, lapply(.SD, function(x) signif(wilcox.test(x ~ INCIDENT_DIAB_T2)$p.value, digits = 2)), .SDcols = table_variables]))
for (i in 1:length(table_variables2)) {
	p_values_cont[length(table_variables)+i] <- signif(fisher.test(rbind(table(table_data[table_data$INCIDENT_DIAB_T2 == 1,][[table_variables2[i]]]), table(table_data[table_data$INCIDENT_DIAB_T2 == 0, ][[table_variables2[i]]])))$p.value, digits = 2)
	}
table1$p_value <- p_values_cont
table1[col == "MEN", col := "WOMEN"] #change the name of the "MEN" variable to "WOMEN" to reflect the inversion
table1 <- rbind(list("N", nrow(table_data), nrow(table_data[table_data$INCIDENT_DIAB_T2 == 1,]), nrow(table_data[table_data$INCIDENT_DIAB_T2 == 0,])), table1, fill=T)
colnames(table1) <- c("Variable", "Total", "With Incident T2D", "Without Incident T2D", "P-value")

table_variables3 <- c("CURR_SMOKE", "MEN", "INCIDENT_DIAB_T2")
table2 <- transpose(merge(table_data[, lapply(.SD, function(x) paste0(round(mean(x, na.rm=T),1), "+-", round(sd(x, na.rm=T),1))), by=EAST, .SDcols=table_variables],
	table_data[, lapply(.SD, function(x) paste0(table(x)[2], " (", round(table(x)[2]/length(x)*100, 1), ")")), by=EAST, .SDcols=table_variables3], by = "EAST"), keep.names = "col", make.names = "EAST")
table2 <- table2[,c("col", "1", "0")]
p_values_cont2 <- as.vector(as.matrix(table_data[, lapply(.SD, function(x) signif(wilcox.test(x ~ EAST)$p.value, digits = 2)), .SDcols = table_variables]))
for (i in 1:length(table_variables3)) {
	p_values_cont2[length(table_variables)+i] <- signif(fisher.test(rbind(table(table_data[table_data$EAST == 1,][[table_variables3[i]]]), table(table_data[table_data$EAST == 0, ][[table_variables3[i]]])))$p.value, digits = 2)
	}
table2$p_value <- p_values_cont2
table2[col == "MEN", col := "WOMEN"] #change the name of the "MEN" variable to "WOMEN" to reflect the inversion
table2 <- rbind(list("N", nrow(table_data[table_data$EAST == 1,]), nrow(table_data[table_data$EAST == 0,])), table2, fill=T)
colnames(table2) <- c("Variable", "From Eastern Finland", "From Western Finland", "P-value")
table_out <- merge(table1, table2, by = "Variable", all = TRUE)
table_out <- table_out[c("N", "WOMEN", "EAST", "INCIDENT_DIAB_T2", table_variables, "CURR_SMOKE"),]
write.csv(table_out, "microbiome_predicts_incident_T2D/Table1.csv", row.names=F)

#Correlations and clustering between the associated taxa in west data
otu_table_assoc_taxa_west <- as.data.frame(otu_table(prune_taxa(ncbi_cox_west$neat_results$Predictor, ncbi_data_raw_west)))
rownames(otu_table_assoc_taxa_west) <- ncbi_cox_results_west$Species[match(rownames(otu_table_assoc_taxa_west), ncbi_cox_results_west$Predictor)]
set.seed(11235)
proprmatrix_west <- propr(t(otu_table_assoc_taxa_west), metric = "rho", p = 100)
clusters_assoc_west <- hclust(dist(proprmatrix_west@matrix), method = "ward.D2")
#Compute the Kelley-Gardner-Sutcliffe penalty function for a hierarchical cluster tree, to determine optimal number of clusters
op_k_west <- kgs(clusters_assoc_west, dist(proprmatrix_west@matrix), maxclus = 20)
op_k_west <- as.numeric(names(op_k_west[which(op_k_west == min(op_k_west))]))
cluster_ids_west <- cutree(tree = clusters_assoc_west, k = op_k_west)
svg("microbiome_predicts_incident_T2D/clusters_west.svg", width=10, height=10)
plot(clusters_assoc_west)
rect.hclust(clusters_assoc_west, k = op_k_west, border = 2:7)
dev.off()

#plot heatmap of taxa associations, clustering, and hazard ratios in the west data
newnames_west <- lapply(rownames(proprmatrix_west@matrix),function(x) bquote(italic(.(x))))

#clusters are identical in membership of taxa in the same cluster, so we can just copy the cluster annotation from east data to west data to keep cluster colors and naming consistent
heatmap_annotation_west <- heatmap_annotation
#get correct hazard ratios for west data
heatmap_annotation_west$HR <- gsub("([0-9]\\.[0-9]*)[[:space:]].*", "\\1", ncbi_cox_results_west$HR[match(heatmap_annotation_west $Predictor, ncbi_cox_results_west$Predictor)])
heatmap_annotation_west$HR <- round(as.numeric(as.character(heatmap_annotation_west $HR)), 1)
heatmap_annotation_west$HR <- factor(heatmap_annotation_west $HR, levels = rev(seq(0.8, 1.2, 0.1)))

svg("microbiome_predicts_incident_T2D/correlations_west.svg", width=15, height=15)
pheatmap(proprmatrix_west@matrix, labels_row = as.expression(newnames_west), labels_col = as.expression(newnames_west), annotation_row = heatmap_annotation_west[4], treeheight_row = 0, annotation_col = heatmap_annotation_west[2], annotation_colors = ann_colors, cutree_rows = op_k, cutree_cols = op_k, clustering_method = "ward.D2", color = heatmap_colors, breaks = seq(-1, 1, length.out = 11), legend_breaks = seq(-1, 1, length.out = 11), cellwidth=10, cellheight=10)
dev.off()

save.image("microbiome_predicts_incident_T2D/Analysis.RData")
```
