## Supplementary material for "Gut microbiome composition is predictive of incident type 2 diabetes": Supplemetary_information.docx

**Supplementary information**

**Table S1.** Results of multivariable adjusted Cox regression models for incident type 2 diabetes.

|  | **Eastern Finland** | | | | **Western Finland** | | |
| --- | --- | --- | --- | --- | --- | --- | --- |
| **Predictor** | Coefficient | Hazard ratio | P-value | Adjusted  p-value | Coefficient | Hazard ratio | P-value |
| PC1 | -0.246 | 0.782 (95% CI, 0.693-0.883) | 1 × 10^-4^ | 0.0099 | -0.067 | 0.935 (95% CI, 0.785-1.114) | 0.4542 |
| Cluster 1 | 0.182 | 1.2 (95% CI, 1.082-1.33) | 5 × 10^-4^ | - | 0.168 | 1.183 (95% CI, 1.018-1.375) | **0.0284** |
| Cluster 2 | 0.123 | 1.13 (95% CI, 1.008-1.267) | 0.0358 | - | 0.062 | 1.064 (95% CI, 0.898-1.261) | 0.4742 |
| Cluster 3 | 0.211 | 1.235 (95% CI, 1.106-1.381) | 2 × 10^-4^ | - | 0.08 | 1.083 (95% CI, 0.917-1.278) | 0.3465 |
| Cluster 4 | 0.217 | 1.242 (95% CI, 1.115-1.384) | 1 × 10^-4^ | - | 0.099 | 1.104 (95% CI, 0.939-1.299) | 0.2306 |
| Cluster 5 | 0.19 | 1.209 (95% CI, 1.092-1.339) | 3 × 10^-4^ | - | 0.163 | 1.177 (95% CI, 1.016-1.364) | **0.0297** |
| *Blautia sp. KLE 1732* | 0.198 | 1.219 (95% CI, 1.09-1.364) | 5 × 10^-4^ | 0.0153 | 0.123 | 1.13 (95% CI, 0.958-1.334) | 0.1469 |
| *Clostridium phoceensis* | 0.197 | 1.218 (95% CI, 1.09-1.361) | 5 × 10^-4^ | 0.0153 | 0.014 | 1.014 (95% CI, 0.858-1.198) | 0.8702 |
| *Coprococcus comes* | 0.194 | 1.214 (95% CI, 1.084-1.36) | 8 × 10^-4^ | 0.0153 | -0.044 | 0.957 (95% CI, 0.807-1.135) | 0.6148 |
| *[Clostridium] citroniae* | 0.192 | 1.212 (95% CI, 1.086-1.352) | 6 × 10^-4^ | 0.0153 | 0.192 | 1.212 (95% CI, 1.035-1.42) | **0.0172** |
| *Eggerthella lenta* | 0.187 | 1.205 (95% CI, 1.08-1.344) | 8 × 10^-4^ | 0.0153 | 0.071 | 1.074 (95% CI, 0.909-1.269) | 0.4022 |
| *Blautia obeum* | 0.18 | 1.197 (95% CI, 1.071-1.338) | 0.0015 | 0.0221 | 0.112 | 1.118 (95% CI, 0.952-1.313) | 0.1747 |
| *[Clostridium] glycyrrhizinilyticum* | 0.176 | 1.193 (95% CI, 1.08-1.318) | 5 × 10^-4^ | 0.0153 | 0.103 | 1.108 (95% CI, 0.942-1.304) | 0.2163 |
| *Anaerostipes hadrus* | 0.171 | 1.186 (95% CI, 1.061-1.327) | 0.0028 | 0.0279 | -0.065 | 0.937 (95% CI, 0.795-1.105) | 0.4416 |
| *[Clostridium] symbiosum* | 0.169 | 1.184 (95% CI, 1.061-1.321) | 0.0025 | 0.0277 | 0.11 | 1.117 (95% CI, 0.949-1.314) | 0.1832 |
| *Bacteroides vulgatus* | 0.166 | 1.18 (95% CI, 1.047-1.331) | 0.0067 | 0.0461 | 0.121 | 1.128 (95% CI, 0.945-1.347) | 0.1821 |
| *Oscillibacter sp.  KLE 1745* | 0.165 | 1.179 (95% CI, 1.054-1.319) | 0.0039 | 0.0325 | 0.091 | 1.096 (95% CI, 0.926-1.296) | 0.2875 |
| *[Clostridium] bolteae* | 0.164 | 1.178 (95% CI, 1.068-1.3) | 0.0011 | 0.0172 | 0.185 | 1.204 (95% CI, 1.043-1.39) | **0.0115** |
| *[Ruminococcus] gnavus* | 0.162 | 1.176 (95% CI, 1.06-1.304) | 0.0023 | 0.0277 | 0.155 | 1.168 (95% CI, 1.006-1.355) | **0.0414** |
| *Eubacterium ramulus* | 0.157 | 1.17 (95% CI, 1.048-1.306) | 0.0053 | 0.0389 | 0.042 | 1.043 (95% CI, 0.885-1.229) | 0.6161 |
| *Tyzzerella nexilis* | 0.15 | 1.162 (95% CI, 1.046-1.292) | 0.0052 | 0.0389 | 0.158 | 1.171 (95% CI, 1.011-1.355) | **0.0347** |
| *Alistipes putredinis* | -0.159 | 0.853 (95% CI, 0.767-0.949) | 0.0035 | 0.0325 | -0.054 | 0.948 (95% CI, 0.805-1.116) | 0.5192 |
| *Sutterella wadsworthensis* | -0.17 | 0.844 (95% CI, 0.752-0.947) | 0.004 | 0.0325 | -0.023 | 0.977 (95% CI, 0.823-1.16) | 0.7939 |
| *Alistipes indistinctus* | -0.177 | 0.838 (95% CI, 0.747-0.94) | 0.0025 | 0.0277 | 0.023 | 1.023 (95% CI, 0.862-1.213) | 0.7961 |

- Significant p-values (< 0.05) for models using data from Western Finland are indicated in bold font.
